# Investigating the Relationship Between Rare Genetic Variants and Fibrosis in Pediatric Nonalcoholic Fatty Liver Disease

**DOI:** 10.1101/2024.03.02.24303632

**Authors:** Julia Wattacheril, Sarah E. Kleinstein, Patrick R. Shea, Laura A. Wilson, G. Mani Subramanian, Robert P. Myers, Jay Lefkowitch, Cynthia Behling, Stavra A. Xanthakos, David B. Goldstein, the NASH Clinical Research Network

**Affiliations:** Columbia University Vagelos College of Physicians and Surgeons, Department of Medicine, Center for Liver Disease and Transplantation, New York Presbyterian Hospital; Columbia University Vagelos College of Physicians and Surgeons, Institute for Genomic Medicine; Department of Epidemiology, Johns Hopkins University; OrsoBio, Inc; Columbia University Vagelos College of Physicians and Surgeons, Department of Pathology; University of California San Diego, Department of Pathology; Department of Pediatrics, University of Cincinnati, Cincinnati Children’s Hospital Medical Center

## Abstract

**Background and Aims:** Nonalcoholic Fatty Liver Disease (NAFLD) is a complex human disease. Common genetic variation in the patatin-like phospholipase domain containing 3 (*PNPLA3*) and transmembrane 6 superfamily member 2 (*TM6SF2*) genes have been associated with an increased risk of developing NAFLD, nonalcoholic steatohepatitis (NASH), and fibrosis in adults. The role of rare genetic variants in the development and progression of NAFLD in children is not well known. We aimed to explore the role of rare genetic variants in pediatric patients with advanced fibrosis.

**Methods:** Whole exome sequencing data was generated for 229 pediatric patients diagnosed with NAFLD recruited from the NASH Clinical Research Network (NASH CRN). Case-control single variant and gene-based collapsing analyses were used to test for rare variants that were enriched or depleted within the pediatric NAFLD cohort specifically for advanced fibrosis (cases) versus those without fibrosis (controls) or six other histologic characteristics. Exome data from non-NAFLD population controls were also used for additional analyses. All results were adjusted for multiple testing using a Bonferroni correction.

**Results:** No genome-wide significant associations were found between rare variation and presence of advanced fibrosis or NASH, nor the severity of steatosis, inflammation, or hepatocellular ballooning. Significantly, no enrichment of rare variants in *PNPLA3* or *TM6SF2* was observed across phenotypes.

**Conclusion:** In a cohort of children with histologically proven NAFLD, no genome-wide significant associations were found between rare genetic variation and advanced fibrosis or six other histologic features. Of particular interest was the lack of association with genes of interest in adults: *PNPLA3* and *TM6SF2,* though limitations in sample size may reduce the ability to detect associations, particularly with rare variation.

## Introduction

Nonalcoholic fatty liver disease (NAFLD) is a common, complex disease with a significant public health impact, affecting more than 25% of the developed world (1). The development of NAFLD, which is strongly associated with obesity, often occurs during childhood and adolescence (2). The inflammatory phenotype of NAFLD, nonalcoholic steatohepatitis (NASH), is an area of urgent interest, given its potential progression to end stage liver disease including cirrhosis, hepatocellular carcinoma, and decompensated disease, which can lead to liver transplant or death. More advanced fibrosis in NASH is strongly associated with these clinical outcomes (3) and earlier onset of advanced disease has been observed with pediatric NAFLD (4) including the need for transplantation (5, 6). Few genetic studies have been conducted in NAFLD and NASH based on liver histology, especially in children and adolescents (7).

Evidence of substantial heritability for NAFLD and NASH is available from epidemiological, familial and twin studies (8–11). Histologic phenotyping allows for accurate fibrosis staging, a relevant outcome in NAFLD. Given the invasiveness of liver biopsy, histologic phenotyping has predominantly been limited to adults. Several genome-wide association studies (GWAS) have discovered and validated an association between the rs738409 (I148M) variant in the Patatin-Like Phospholipase Domain Containing 3 (*PNPLA3*) *gene,* encoding adiponutrin, the major genetic risk factor for NAFLD (12). PNPLA3 is a transmembrane protein with diacyl and triacylglycerol lipase activity that is expressed in liver and adipose tissue (13). The *PNPLA3* I148M variant is associated with increased risk of hepatic steatosis, elevated transaminase levels, and is notable for its influence on the risk for both NASH and fibrosis (14, 15). Four other single nucleotide polymorphisms (SNPs) have been associated with NAFLD histologic phenotypic features. The biological processes involved with these SNPs include regulation of cell adhesion/lipoprotein metabolism (*NCAN,* neurocan*/TM6SF2*), glucose metabolism (*GCKR,* glucokinase regulator) and triglyceride catabolism (*LYPLAL1,* lysophospholipase like 1)(16). Other studies have found the rs641738 C>T variant in membrane bound O-acyltransferase domain containing 7 (*MBOAT7*), which encodes a protein involved in phospholipid remodeling, is associated with histologic endpoints of inflammation and fibrosis (17, 18).

Advanced disease at an early age, confirmed with histopathology, represents an extreme phenotype of particular clinical interest and with potentially greater genetic risk. Under this circumstance, the greater impact of rare variants may influence risk of fibrotic disease more than the common variants seen with GWAS (19). Previously, in a cohort of 223 pediatric patients with NAFLD, no association was found between any common SNP in *PNPLA3* and histologic parameters (20). Genetic variants in genes regulating insulin receptor activity (ectonucleotide pyrophosphatase/phosphodiesterase 1 (*ENPP1),* insulin receptor substrate 1 (*IRS1)*) and antioxidant activity (mitochondrial superoxide dismutase 2 (*SOD2)*) have been associated with pediatric fibrosing NASH (21, 22); the *TM6SF2* rs559542986 minor allele has also been associated with ≥ stage 1 fibrosis in pediatric NAFLD (23). Regulation of Kruppel-like factor 6 (*KLF6*) splicing by the rs3750861 intronic SNP has also been associated with fibrogenesis in pediatric NASH (24). Homozygosity for the rs13412852 variant of Lipin-1 (*LPIN1*) was associated with the absence of hepatic fibrosis in children with NAFLD, independent of *PNPLA3* genotype (25). A recent small study revealed evidence for variants in *GCKR* and uncoupling protein 2 (*UCP2*) associated with fibrosing NAFLD in adolescents (26).

The role of rare variants in common complex disease is gaining attention due to the recent application of next-generation sequencing in disease genetics studies, including in liver diseases. A large-scale whole-exome sequencing (WES) study identified loci associated with serum aminotransferase levels. A common splice variant in hydroxysteroid 17-beta dehydrogenase (*HSD17B13)* was reported to be associated with reduced levels of aminotransferases, as well as protection against both alcoholic and nonalcoholic cirrhosis (27); validation included a pediatric cohort of Hispanic American children and adolescents. More recently, a missense variant in mitochondrial amidoxime reducing component 1 (*MARC1)* was found to associate with protection from cirrhosis, as well as with reduced levels of liver enzymes and cholesterol (28). The associations with reduced risk observed for the *HSD17B13* rs72613567 splice variant and *MARC1* A165T substitution have been subsequently replicated in an independent pediatric NAFLD cohort (29).

The potential application of next-generation sequencing in a pediatric NAFLD population is ideal; children and adolescents lack many of the environmental stressors that confound studies of adult NAFLD, such as alcohol, medications, and co-morbid illnesses. Because some children with NAFLD display advanced fibrosis at an early age, an extreme phenotype which may represent exceptional genetic vulnerability, we sought to identify causal variants associated with advanced fibrosis using WES. Simultaneously, given the well-phenotyped subjects in the Nonalcoholic Steatohepatitis Clinical Research Network (NASH CRN), we sought to directly interrogate associations with other histologic components (inflammation, steatosis, ballooning injury) previously described in this cohort (7), as opposed to testing serum-based or anthropometric associations previously undertaken in non-histologically phenotyped cohorts (8, 30).

## Materials and Methods

All pediatric participants enrolled in the NASH CRN in pediatric clinical trials with *advanced fibrosis*, stage 3 or 4 fibrosis (n=116) were eligible for inclusion. Subjects were selected based on fibrosis scores and available DNA. A total of 114 pediatric NAFLD controls *without any fibrosis* (stage 0) were selected based upon DNA availability and matching for age, sex and body mass index (BMI) Z-score. Clinical and histologic features of pediatric database participants have been described (31). Subjects met exclusion criteria for any other potential contributors to fatty liver disease. All protocols for NASH CRN studies were approved by Institutional Review Boards at each participating center, the Data Coordinating Center, and a Data Safety and Monitoring Board appointed by the National Institutes of Health and were conducted in accordance with both the declarations of Helsinki and Istanbul. All parents or guardians provided consent for their child’s participation and all children >7 years of age provided assent.

Liver biopsies from pediatric patients diagnosed with NAFLD in the NASH CRN database were reviewed by the NASH CRN Pathology Committee during quarterly meetings at a multiheaded microscope. Consensus scores for a variety of histologic parameters were assigned based on pre-determined histologic criteria (32). At the time of review, the pathologists were unaware of clinical or demographic features of any subject. Key histologic components of NASH were evaluated as follows: Steatosis (% of hepatocytes containing a fat droplet): 0 (none to <5%), 1 (5% to 33%), 2 (34% to 66%), and 3 (>66%). Lobular inflammation assessment was based on the number of inflammatory foci per 200x field averaged over the entire biopsy and scored 0= no foci,1: <2 foci per 200x field,2: 2-4 foci per 200x field, and 3: >4 foci per 200x. Portal inflammation was categorized as none, mild, and more than mild. Ballooning was scored as: none, few, and many. Fibrosis stage was assigned as follows: stage 0 (no fibrosis), stage 1a (mild zone 3 perisinusoidal requiring trichrome stain), stage 1b (moderate zone 3 perisinusoidal fibrosis visible on H&E), stage 1c (portal/periportal fibrosis only), stage 2 (zone 3 perisinusoidal and periportal), stage 3 (bridging fibrosis), and stage 4 (cirrhosis).

A diagnostic category for each biopsy was assigned based on the constellation of typical features of NASH in the biopsy without regard to NAFLD Activity Score (NAS). The NAS is the sum of the scores assigned for steatosis, lobular inflammation and ballooning, as described (32). Diagnoses include: NAFLD not NASH, borderline zone 1 NASH (portal pediatric Type 2 pattern), borderline zone 3 NASH (central adult Type 1 pattern), and definite NASH (33). These diagnostic categories were grouped categorically for analyses.

WES was performed on 230 pediatric NAFLD (n=114 advanced fibrosis, n=116 controls) patients of all ethnicities using Illumina HiSeq2500 DNA sequencers using standard protocols. Briefly, sequences were aligned to the Human Reference Genome (NCBI Build 37/hg19) using the Burrows-Wheeler Alignment Tool (BWA) v0.5.10 (34). Picard v1.59 software (Broad Institute, Boston, MA; http://broadinstitute.github.io/picard/) was used to remove duplicate reads and generate BAM files. We recalibrated base quality scores, performed a local realignment of indels, and called variants using Genome Analysis Toolkit (GATK) v1.6 (35). Variants were annotated to Ensembl 73 using SnpEff v3.3. Following alignment, we performed standard quality control measures to exclude contaminated, low coverage, related, or gender discordant samples. Following quality control and the exclusion of one patient with a genetic diagnosis and clinical confirmation of Wilson Disease (36), the final cohort for analysis consisted of 229 unrelated pediatric NAFLD patients with high quality sequences (≥40-fold average coverage). In addition, exome sequence data obtained from 11,295 subjects enrolled in other studies unrelated to liver or fibrotic diseases were utilized as population controls for certain analyses. These samples underwent identical bioinformatic processing and quality control.

An analysis of genetic ancestry was performed using the EIGENSTRAT software package (37) using common SNPs from the sequencing dataset. Principal components (PCs) were calculated independently for the 229 pediatric patients (cases with advanced fibrosis and matched controls from the NASH CRN) as well as for the combined pediatric cases and population controls (**Supplementary Figure 1**). Association analyses were performed using the Analysis Tool for Annotated Variants (ATAV) v6.6 software package (https://github.com/igm-team/atav/) (38).

Case-control single variant and gene-based collapsing analyses were used to test for rare variants that were enriched or depleted within the pediatric NAFLD cohort across 1) the primary endpoint: advanced fibrosis (115 bridging/cirrhotic stage 3/4 vs. 114 none (stage 0)), 2) other histologic features with clinical relevance: steatosis (160 scored 2/3 vs. 69 scored 0/1), lobular inflammation (101 scored 2/3 vs. 128 scored 0/1), portal inflammation (202 scored as mild/more than mild vs. 26 scored as none; one unable to be scored), hepatocyte ballooning (86 scored as few/many vs. 143 none), NASH (132 aggregate definite and borderline vs. 97 no NASH and NAFLD activity score (NAS; 96 scored >4 vs. 133 scored ≤4). Histologic features other than fibrosis were categorized based on severity of the individual feature: minimal steatosis and lobular inflammation were grouped with none given the minor features required to qualify as a score of 1 in order to enhance detection of associations with moderate to severe phenotypes. Additional analyses compared the pediatric NAFLD cohort to previously sequenced population controls. Single-variant analyses were performed to investigate 1) rare genetic variation using a Fisher’s exact test (FET) of functional variants with a minor allele frequency (MAF<5%; and 2) common (≥5% MAF), functional variation using a logistic regression model adjusted for age (pediatric-only analyses), gender, and ancestry. For gene-based collapsing analyses, six qualifying variant frameworks (39) were defined using separate MAF thresholds [Exome Aggregation Consortium (ExAC) MAF=1%, 0.05% (rare), or 0% (ultra-rare)] and variant annotation [loss-of-function (LoF), damaging, or non-synonymous] (**Supplementary Table 1**). For gene-based analysis, a synonymous variant model was used as a negative control. Thresholds for statistical significance were determined using a Bonferroni correction to adjust for the number of variants or genes in each analysis (p<3.03E-6 to correct for 16,525 common functional variants or p<2.68E-6 to correct for 18,652 genes). All authors had access to the study data and reviewed and approved the final manuscript.

## Results

Clinical and laboratory characteristics of the 229 NAFLD subjects included are shown in **Table 1**. The median age was 12.9 years (SD 2.5). Mean BMI, BMI Z-score, and serum aminotransferase values were elevated and consistent with that reported previously for pediatric NAFLD patients. With regard to diagnostic histologic patterns comprising the extremes of phenotype of interest, 115 cases had stage 3 or 4 fibrosis and 114 had no fibrosis. Overall, 132 subjects had a definite or borderline NASH including 109 of the 115 with advanced fibrosis. The median overall NAS was 4.2 (SD 1.5).

**Table 1.** Baseline Clinical Characteristics, N= 229.

| Characteristic | Mean (SD) |
| --- | --- |
| **Demographic Characteristics** |  |
| Age (years) | 12.9 (2.5) |
| Height (cm) | 159.2 (13.3) |
| Weight (kg) | 82.3 (24.5) |
| BMI (kg/m<sup>2</sup>) | 31.8 (6.3) |
| BMI Z Score | 2.21 (0.42) |
| **Laboratory Measures** |  |
| ALT (U/L) | 115.6 (101.3) |
| AST (U/L) | 72.2 (61.8) |
| **Histologic Characteristics** |  |
| **Fibrosis stage** | **N (%)** |
| 0 | 114 (49.8) |
| 3 | 100 (43.7) |
| 4 | 15 (6.5) |
| **Steatosis** | **N (%)** |
| 0 | 3 (1.3) |
| 1 | 66 (28.8) |
| 2 | 59 (25.8) |
| 3 | 101 (44.1) |
| **Lobular inflammation** | **N (%)** |
| 0 | 0 (0) |
| 1 | 128 (55.9) |
| 2 | 81 (35.4) |
| 3 | 20 (8.7) |
| **Portal inflammation** | **N (%)** |
| 0 | 26 (11.4) |
| 1 | 148 (64.9) |
| 2 | 54 (23.7) |
| **Ballooning** | **N (%)** |
| 0 | 143 (62.5) |
| 1 | 53 (23.1) |
| 2 | 33 (14.4) |
| **NASH** | **N (%)** |
| Definite | 52 (22.7) |
| Borderline | 80 (34.9) |
| No NASH | 97 (42.4) |
|  | **Mean (SD)** |
| **NAFLD Activity Score** | 4.17 (1.52) |

No genome-wide significant associations were found between rare variation and advanced fibrosis (**Table 2**, **Figure 1A-B**), presence of histologic NASH (**Table 3**, **Figure 1C-D**), or scores for steatosis, lobular inflammation, hepatocyte ballooning, portal inflammation, or total NAS within the pediatric NAFLD cohort. Additionally, there were no genome-wide significant associations observed with rare variants when the pediatric NAFLD cohort was compared to a large, previously sequenced population control cohort. No enrichment of rare variants in *PNPLA3* or *TM6SF2* was observed across clinical phenotypes.

**Table 2.** Fibrosis [115 fibrosis stage 3/4 (cases) vs 114 stage 0 (controls)] gene-based collapsing analysis: top, non significant hits enriched in the rare, damaging dominant model.

| Rank | Gene Name | Case Carriers | Frequency of Carriers among Cases | Control Carriers | Frequency of Carriers among Controls | Enrichment Direction | p-value |
| --- | --- | --- | --- | --- | --- | --- | --- |
| 1 | *NLRP13* | 0 | 0 | 5 | 0.04 | control | 0.03 |
| 2 | *MACF1* | 5 | 0.04 | 0 | 0 | case | 0.06 |
| 3 | *PML* | 4 | 0.03 | 0 | 0 | case | 0.12 |
| 4 | *MYLK* | 4 | 0.03 | 0 | 0 | case | 0.12 |
| 5 | *SVIL* | 0 | 0 | 3 | 0.03 | control | 0.12 |
| 6 | *HTT* | 0 | 0 | 3 | 0.03 | control | 0.12 |
| 7 | *USH1C* | 0 | 0 | 3 | 0.03 | control | 0.12 |
| 8 | *DNAH10* | 0 | 0 | 3 | 0.03 | control | 0.12 |
| 9 | *TBXAS1* | 0 | 0 | 3 | 0.03 | control | 0.12 |
| 10 | *HERC1* | 0 | 0 | 3 | 0.03 | control | 0.12 |

Highlighted (yellow) genes were also present among the top results of the other rare/damaging collapsing models.

**Figure 1A.**
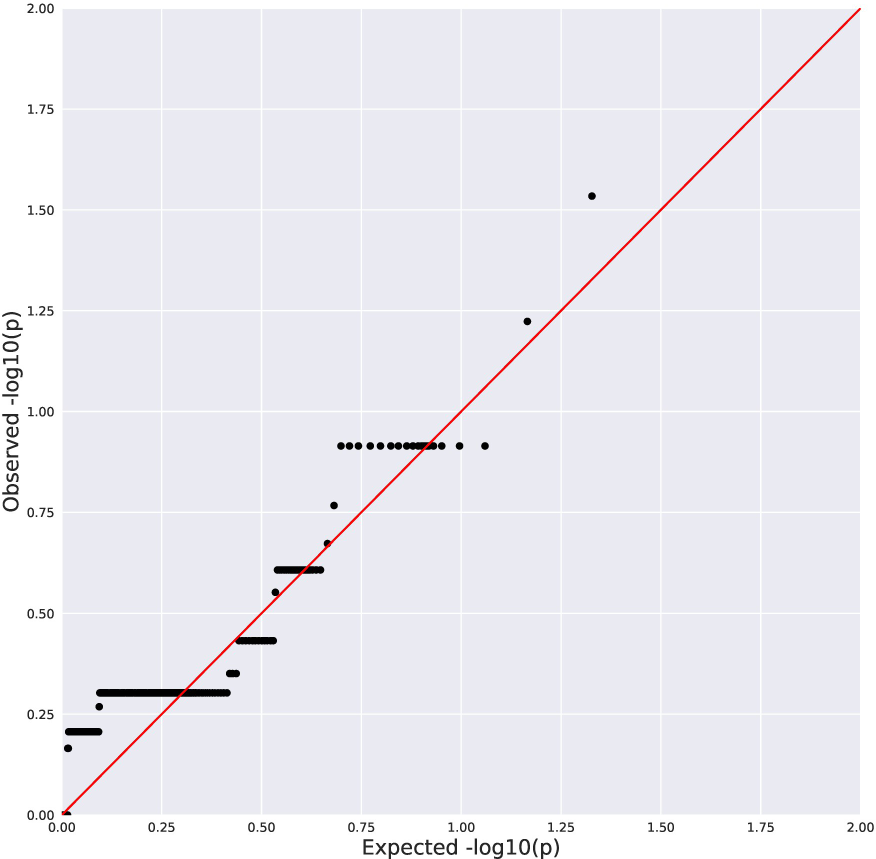
Quantile-Quantile Plot of Collapsing Analysis Results for Stage 3/4 Fibrosis vs Stage 0 (rare, damaging dominant genetic model). Black dots indicate the observed P value for a gene plotted against a corresponding quantile from the null distribution. Red line indicates expected P values if the observed results were consistent with the null hypothesis. Genomic inflation factor (λ)=1.01.

**Figure 1B.**
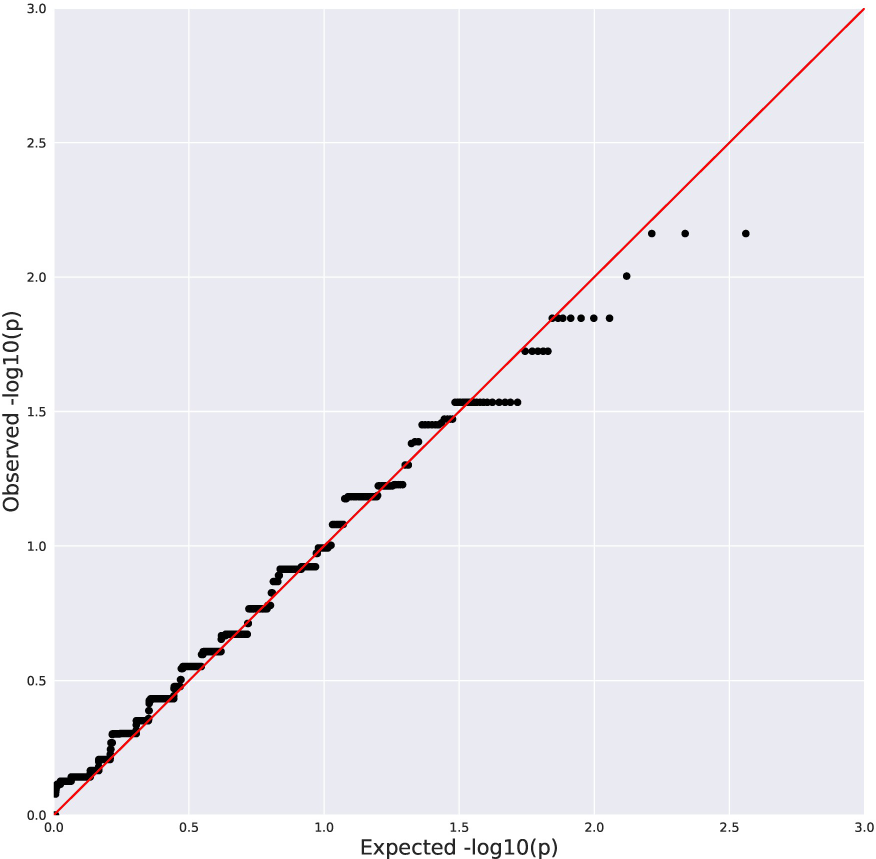
Quantile-Quantile Plot of Collapsing Analysis Results for Stage 3/4 Fibrosis vs Stage 0 (synonymous negative control model). Black dots indicate the observed P value for a gene plotted against a corresponding quantile from the null distribution. Red line indicates expected P values if the observed results were consistent with the null hypothesis. λ=1.00.

**Figure 1C.**
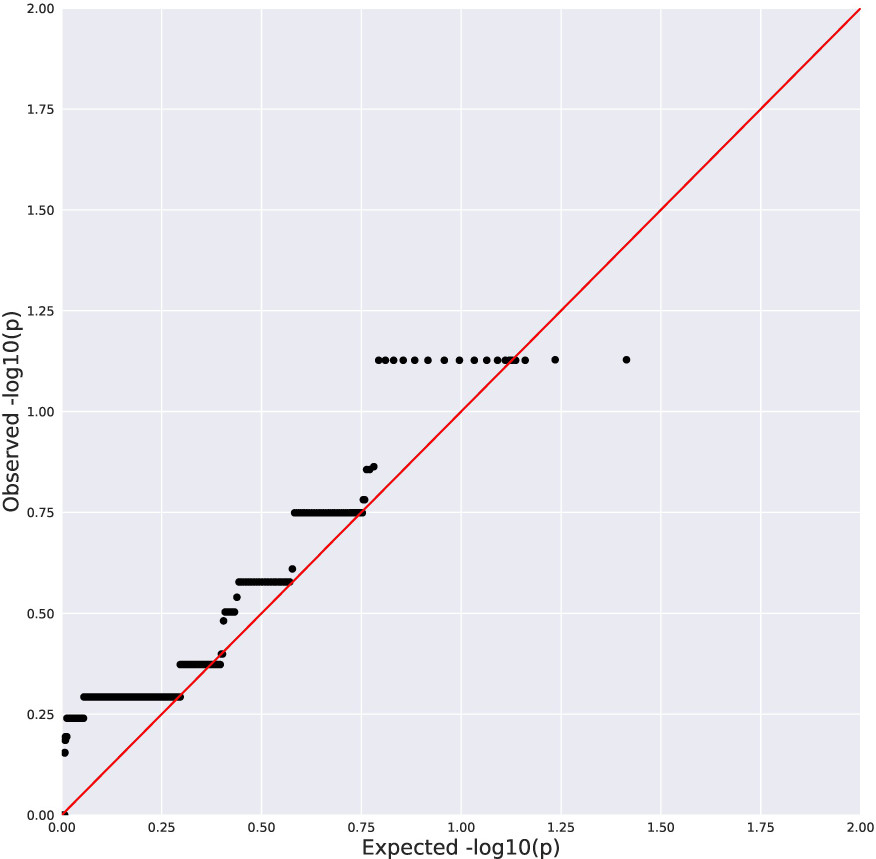
Quantile-Quantile Plot of Collapsing Analysis Results for NASH vs No NASH (rare, damaging dominant genetic model). Black dots indicate the observed P value for a gene plotted against a corresponding quantile from the null distribution. Red line indicates expected P values if the observed results were consistent with the null hypothesis. λ=1.04.

**Figure 1D.**
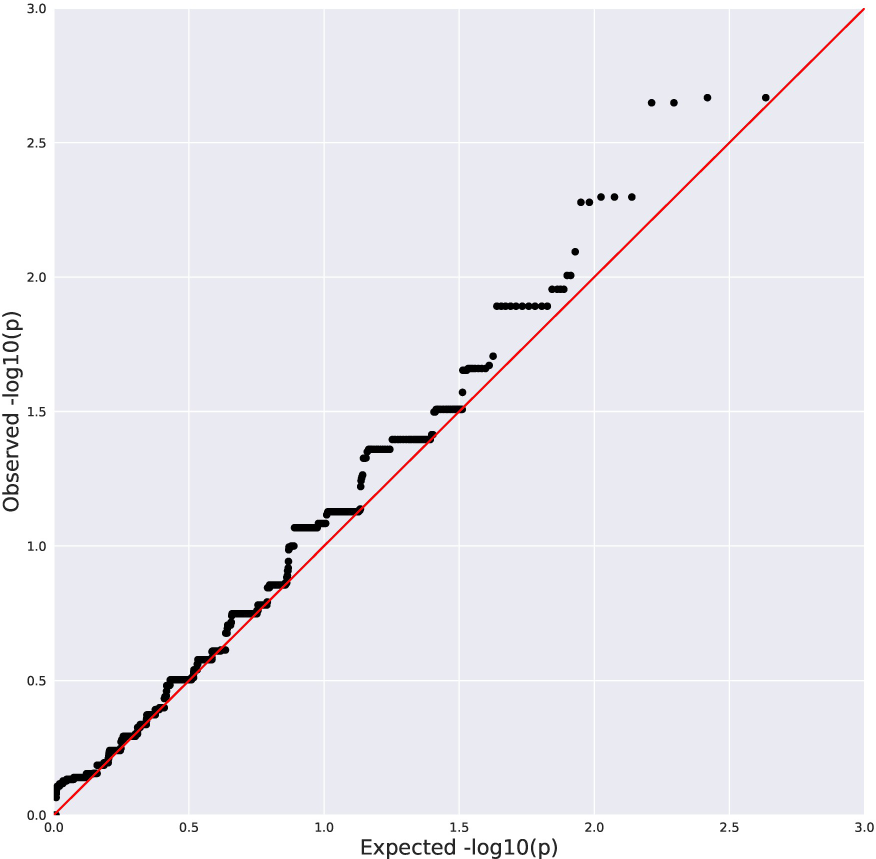
Quantile-Quantile Plot of the Results from Collapsing Analysis of NASH vs No NASH (synonymous negative control model). Black dots indicate the observed P value for a gene plotted against a corresponding quantile from the null distribution. Red line indicates expected P values if the observed results were consistent with the null hypothesis. λ=1.04.

**Table 3.** NASH [132 aggregate NASH (cases) vs 97 no NASH (controls)] gene-based collapsing analysis: top, nonsignificant hits enriched in the rare, damaging dominant model.

| Rank | Gene Name | Case Carriers | Frequency of Carriers among Cases | Control Carriers | Frequency of Carriers among Controls | Enrichment Direction | p-value |
| --- | --- | --- | --- | --- | --- | --- | --- |
| 1 | *PKHD1* | 5 | 0.04 | 0 | 0 | case | 0.07 |
| 2 | *MACF1* | 5 | 0.04 | 0 | 0 | case | 0.07 |
| 3 | *HTT* | 0 | 0 | 3 | 0.03 | control | 0.08 |
| 4 | *USH1C* | 0 | 0 | 3 | 0.03 | control | 0.08 |
| 5 | *PREX2* | 0 | 0 | 3 | 0.03 | control | 0.08 |
| 6 | *DNAH10* | 0 | 0 | 3 | 0.03 | control | 0.08 |
| 7 | *HERC1* | 0 | 0 | 3 | 0.03 | control | 0.08 |
| 8 | *TRAPPC9* | 0 | 0 | 3 | 0.03 | control | 0.08 |
| 9 | *TMCC3* | 0 | 0 | 3 | 0.03 | control | 0.08 |
| 10 | *CUX1* | 0 | 0 | 3 | 0.03 | control | 0.08 |

Highlighted (yellow) genes were also present among the top results of the other rare/damaging collapsing models.

While there was no enrichment of rare variants, there was enrichment of the common variants in *PNPLA3* and *TM6SF2*. Detailed inspection of genotypes in the full pediatric cohort compared to population controls indicated a large enrichment of the 148M allele of *PNPLA3* in the pediatric cohort. The *PNPLA3* 148M risk allele had a frequency of 73% in the pediatric NAFLD cohort compared to only 24% among population controls (p=1.52e-35, ancestry corrected logistic regression model) and 26% in the 1000 Genomes Project and ExAC population databases. For *TM6SF2* E167K, the 167K risk allele was present at a frequency of 12% in the pediatric NAFLD cohort, double the 6% among population controls (p=1.92e-9, ancestry corrected logistic regression model) and the 7% frequency observed in population databases. However, unlike previous studies, detailed interrogation of the rs738409 (I148M) SNP and rs58542926 (E167K) SNP did not reveal any significant association with the risk of advanced fibrosis or any other clinical endpoint within the pediatric cohort (**Table 4A**). Further, there were no enrichment of rare, damaging mutations in *PNPLA3* or *TM6SF2* among any clinical phenotype investigated. A detailed comparison of ancestry in the patient groups with advanced or no fibrosis did not indicate any systematic biases in composition of the groups (**Figure 2A-B**). To assess the influence of rare genetic variation in the context of the *PNPLA3* 148M allele, we performed separate case-control analysis stratified by *PNPLA3* carrier status. No statistically significant rare variant associations were detected in collapsing analyses for case and control subjects with one or more *PNPLA3* 148M allele or for those with two copies of the I148 reference allele (**Supplementary Table 2**).

**Table 4A.** **Logistic Regression Analysis of *PNPLA3*/*TM6SF2* Risk Alleles and Histologic Measures of NAFLD.** Additive genetic model, adjusted for gender and ancestry; P values indicate probability of observing the difference in MAF between cases/controls by chance.

| Phenotype | PNPLA3 I148M |  |  | TM6SF2 E167K |  |  |
| --- | --- | --- | --- | --- | --- | --- |
|  | Case AF | Control AF | P (logistic) | Case AF | Control AF | P (logistic) |
| **Fibrosis** | 0.77 | 0.70 | 0.08 | 0.13 | 0.11 | 0.65 |
| **Steatosis** | 0.73 | 0.75 | 0.66 | 0.15 | 0.06 | 0.02 |
| **NASH** | 0.74 | 0.73 | 0.58 | 0.12 | 0.12 | 0.95 |
| **Lobular inflammation** | 0.75 | 0.72 | 0.94 | 0.15 | 0.10 | 0.06 |
| **Hepatocyte ballooning** | 0.72 | 0.74 | 0.99 | 0.15 | 0.10 | 0.28 |
| **Portal inflammation** | 0.75 | 0.56 | 0.01 | 0.12 | 0.15 | 0.34 |
| **NAS** | 0.73 | 0.74 | 0.62 | 0.18 | 0.08 | 0.005 |

**Figure 2A.**
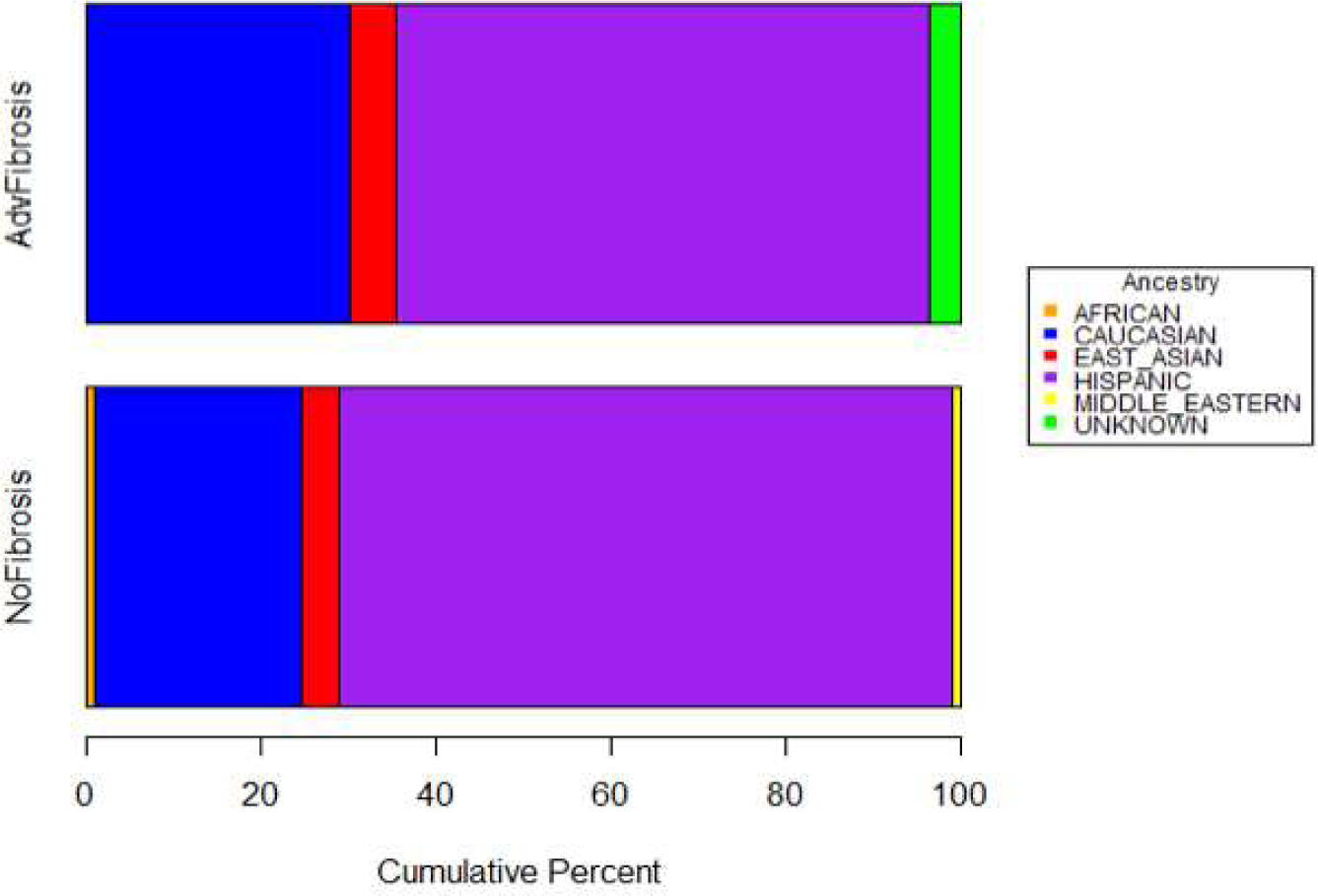
Comparison of Genetically-Inferred Ancestry Among Pediatric Patients with Advanced or No Fibrosis. Overall Distribution of Ancestry Among Fibrosis Groups.

**Figure 2B.**
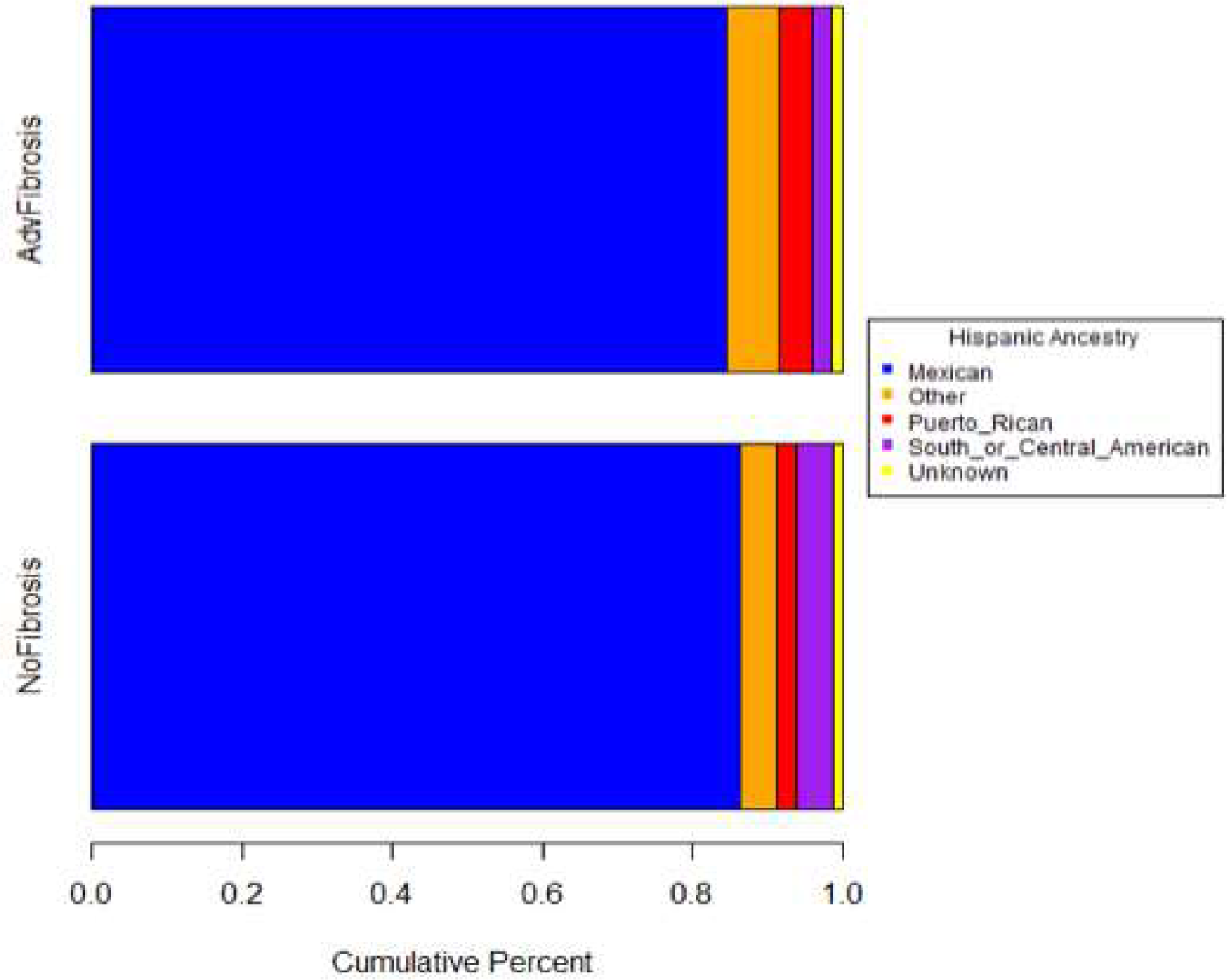
Comparison of Genetically-Inferred Ancestry Among Pediatric Patients with Advanced or No Fibrosis. Comparison of Detailed Self-Reported Ancestries Among Hispanic Subgroup.

A common splice donor variant in the *HSD17B13* gene has been reported to be associated with protection against chronic liver disease in both adult and pediatric patients (27, 40). To examine the influence of *HSD17B13* alleles among pediatric patients enrolled in the NASH CRN, variants computationally predicted to result in frameshifts, premature termination, start/stop loss, or alter splicing of *HSD17B13* were tested for associations with pediatric NAFLD and advanced liver fibrosis. Ancestry corrected logistic regression modeling of the previously implicated rs72613567 splice variant revealed a similar protective trend, with depletion of the minor allele among pediatric NAFLD patients relative to population controls (OR=0.63; p=0.0042), though this level of association is not statistically significant on an exome-wide level. No evidence of association was found between the rs72613567 splice variant and the risk of advanced fibrosis (p=0.65, logistic regression of patients with advanced versus no fibrosis, adjusted for gender and ancestry), with the protective allele at a higher frequency among the advanced fibrosis group (MAF=12.2%) relative to pediatric NAFLD patients with no fibrosis (MAF=10.1%) (**Table 4B**).

**Table 4B.** Logistic Regression Analysis of *HSD17B13* Splice Variant and *MARC1* A165T Risk Alleles and Histologic Measures of NAFLD. Additive genetic model, adjusted for gender and ancestry; P values indicate probability of observing the difference in MAF between cases/controls by chance.

| Phenotype | HSD17B13 Splice |  |  | MARC1 A165T |  |  |
| --- | --- | --- | --- | --- | --- | --- |
|  | Case AF | Control AF | P (logistic) | Case AF | Control AF | P (logistic) |
| **Fibrosis** | 0.12 | 0.10 | 0.65 | 0.15 | 0.18 | 0.29 |
| **Steatosis** | 0.11 | 0.11 | 0.65 | 0.16 | 0.19 | 0.20 |
| **NASH** | 0.12 | 0.10 | 0.62 | 0.16 | 0.17 | 0.68 |
| **Lobular inflammation** | 0.08 | 0.14 | 0.08 | 0.15 | 0.18 | 0.58 |
| **Hepatocyte ballooning** | 0.14 | 0.09 | 0.32 | 0.17 | 0.16 | 0.97 |
| **Portal inflammation** | 0.12 | 0.08 | 0.48 | 0.16 | 0.19 | 0.39 |
| **NAS** | 0.10 | 0.12 | 0.42 | 0.16 | 0.17 | 0.57 |

Recently, a common variant (rs2642438, Ala165Thr) in the *MARC1* gene was reported to be protective against liver disease and cirrhosis (28). We observed a similar depletion of the protective (165Thr) allele among the pediatric NAFLD patients in the NASH CRN cohort relative to population controls (16% vs 26%; **Table 4B**), though this difference in allele frequency was largely accounted for by differences in genetic ancestry (p=1.3E-5 logistic regression with no ancestry correction, p=0.035 after inclusion of ancestry covariates). However, a protective effect (OR=0.75), consistent with that reported by Emdin *et al*, remained after ancestry correction, therefore, as with *HSD17B13*, we cannot rule out a weak protective effect for this variant in our pediatric cohort (28). Though the A165T variant was additionally not found to be associated with any of the clinical phenotypes among our pediatric NAFLD patients (**Table 4B**).

## Discussion

Individuals with advanced fibrosis in NAFLD are a phenotype of interest in adults and children. Understanding the genetic risk variants that contribute to fibrosis development may serve as a therapeutic target. Given early disease onset in children, we interrogated the genetics of NAFLD in a cohort of children with histologically proven NAFLD with advanced fibrosis from the NASH CRN; this cohort represents a unique, carefully characterized and more extreme NAFLD phenotype. WES and detailed interrogation of risk variants within this pediatric cohort did not reveal any significant association between rare variation and the risk of advanced fibrosis or any of seven other unique histologic features. Although we aimed for specific associations with fibrosis, a predictor of fibrosing NAFLD includes the development of steatosis, inflammation and ballooning cell injury *en route* to more advanced disease. Aminotransferase elevations as a surrogate for hepatic fat accumulation and related injury were not interrogated in this cohort, as our primary goal was to investigate direct histologic phenotypes.

Of particular interest within pediatric cases with advanced fibrosis was the lack of enrichment in established genes of interest for fibrosis in adults (14, 41, 42): *PNPLA3* and *TM6SF2*. We did observe enrichment of the known adult risk variants in *PNPLA3* and *TM6SF2* among the entire pediatric cohort (absence and presence of fibrosis) when compared to population controls. In particular, the *PNPLA3* I148M SNP was markedly enriched among the pediatric cohort, emphasizing the critical importance of this factor in pediatric NAFLD susceptibility. This overall enrichment of *PNPLA3* I148M in pediatric NAFLD patients may indicate their high risk for NAFLD and progression overall, which may explain our inability to detect associations with this variant when the pediatric cohort was stratified by histologic endpoints. While the observed association between enrichment in the *PNPLA3* I148M SNP and the absence of association of any unique variant with advanced fibrosis may potentially be attributed to the size of our cohort, our findings are consistent with that from another group specifically investigating the relationship of *PNPLA3* with histologic parameters in pediatric NAFLD (20). Additionally, targeting pediatric steatofibrotic disease (without NASH) may increase the potential for discovering loci specific to fibrosis in pediatric NAFLD with clinical outcomes of interest, namely early onset cirrhosis (43).

In addition to the associations with *PNPLA3* and *TM6SF2*, we also observed a modest protective effect for the common *HSD17B13* splice variant rs72613567 among this pediatric cohort relative to population controls. However, within the pediatric cohort, no protective effect was observed in our analysis of advanced fibrosis. Frequency differences in other *HSD17B13* truncation alleles among Hispanic cases and controls were found to be predominantly driven by population stratification in our cohort and were effectively controlled by a formal correction for genetic ancestry.

Given the growing incidence of NAFLD worldwide, the identification of factors that influence disease risk and progression, particularly among vulnerable populations such as children, is an area of unmet need. It remains unclear whether pediatric NAFLD has the same genetic risk factors as adult NAFLD. Larger, longitudinal studies of children with NAFLD of Hispanic and non-Hispanic ancestry with well-phenotyped histologic features with clinical outcomes are especially needed to conclusively determine whether rare genetic variation is an important factor in pediatric NAFLD with advanced fibrosis. Furthermore, comparisons to large pooled data from adult populations may enhance our understanding of the genomic risk, particularly for fibrosis, across the lifespan of this prevalent, progressive and complex disease.

## Data Availability

All data produced in the present study are available upon reasonable request to the authors.

## ClinicalTrials.gov identifier

NAFLD Pediatric Database 2: NCT01061684, NCT00063635, NCT01529268

## Acknowledgements

The authors thank the National Institute of Diabetes and Digestive and Kidney Diseases (NIDDK) for its support of the NASH CRN and this research. The authors thank the Nonalcoholic Steatohepatitis Clinical Research Network (NASH CRN) investigators and the Ancillary Studies Committee for providing clinical samples and relevant data from the Nonalcoholic Fatty Liver Disease (NAFLD) Pediatric Database 2. The biospecimens from the NASH CRN reported on here were supplied by the NIDDK Central Repository.

## Disclaimer

The content is solely the responsibility of the authors and does not necessarily represent the official views of the National Institutes of Health. This manuscript was not prepared in collaboration with the NIDDK Central Repository and does not necessarily reflect the opinions or official views of the NIDDK Central Repository or the NIDDK.

## Author Contributions

**Julia Wattacheril** study concept and design, acquisition of data; analysis and interpretation of data; drafting of manuscript; critical revision of the manuscript for important intellectual content; study supervision

**Sarah E. Kleinstein** acquisition of data; analysis and interpretation of data; drafting of manuscript; statistical analysis

**Patrick R. Shea** study concept and design; acquisition of data; analysis and interpretation of data; drafting of manuscript; statistical analysis; critical revision of the manuscript for important intellectual content

**Laura A. Wilson** acquisition of data; drafting of manuscript; administrative and material support

**G. Mani Subramanian** critical revision of the manuscript for important intellectual content; obtained funding; material support

**Robert P. Myers** analysis and interpretation of data, critical revision of the manuscript for important intellectual content; obtained funding; material support

**Jay Lefkowitch** acquisition of data

**Cynthia Behling** acquisition of data; analysis and interpretation of data; drafting of manuscript; critical revision of the manuscript for important intellectual content

**Stavra A. Xanthakos** acquisition of data

**David B. Goldstein** study concept and design, acquisition of data; analysis and interpretation of data; critical revision of the manuscript for important intellectual content; study supervision; obtained funding; technical and material support

## *Members of the Nonalcoholic Steatohepatitis Clinical Research Network

### Pediatric Clinical Centers

**Baylor College of Medicine, Houston, TX:** Paula M Hertel, MD; Alberto Ayala Aguilar, MD; Laurel Cavallo, BS; Donna Garner, CPNP; Krupa R Mysore, MD; Alison Shaw, BS; Mary Elizabeth Tessier, MD; Nicole Triggs, CPNP; Cynthia Tsai, BS

**Cincinnati Children’s Hospital Medical Center, Cincinnati, OH:** Stavra Xanthakos, MD; Ana Catalina Arce-Clachar, MD; Kristin Bramlage, MD; Kim Cecil, PhD; Nicole Chaaban, BS; Marialena Mouzaki, MD; Ann Popelar, MPH, CCRP; Andrew Trout, MD

**Emory University, Atlanta, GA:** Miriam Vos, MD, MSPH; Adina Alazraki, MD; Carmen Garcia; Jorge Jara-Garra; Saul Karpen, MD, PhD

**Indiana University School of Medicine/Riley Hospital for Children, Indianapolis, IN:** Jean P. Molleston, MD; Oscar W. Cummings, MD; Kathryn Harlow Adams, MD; Ashley Hartman, CMA; Kelley S. Jackson, RN; Chaowapong Jarasvaraparn, MD; Sandie Kennedy, NP; Ann Klipsch, RN; Wendy Morlan, RN; Emily Ragozzino, CCRC; Kyla Tolliver, MD

**Northwestern University Feinberg School of Medicine/Ann & Robert H. Lurie Children’s Hospital of Chicago:** Mark H. Fishbein, MD; Angela Anthony, BA, CRC; Catherine Chapin, MD

**Saint Louis University, St Louis, MO:** Ajay K. Jain, MD; Danielle Carpenter, MD; Theresa Cattoor, RN; Paige Puricelli, RN

**University of California San Diego, San Diego, CA:** Jeffrey B. Schwimmer, MD; Amy Alba, MPH; Cynthia Behling, MD, PhD; Nidhi Goyal, MD, MPH; Leila Keyvan; Michael S. Middleton, MD, PhD; Rebecca Morfin; Kimberly Newton, MD; Claude Sirlin, MD; Jaret Skonieczny; Patricia Ugalde-Nicalo, MD, MAS; Karenina Valdez, MD; Andrew Wang, MD

**University of California San Francisco, San Francisco, CA:** Ryan Gill, MD, PhD

**University of Washington Medical Center and Seattle Children’s Hospital, Seattle, WA:** Niviann Blondet, MD; Camila Khorrami, BS; Randolph Otto, MD; Matthew Yeh, MD, PhD; Melissa Young, CCRC

### Resource Centers

**National Cancer Institute, Bethesda, MD:** David E. Kleiner, MD, PhD

**Data Coordinating Center, Johns Hopkins University, Bloomberg School of Public Health, Baltimore, MD:** James Tonascia, PhD; Peggy Adamo, BS; Patricia Belt, BS; Jeanne M. Clark, MD, MPH; Jennifer M. DeSanto, RN, BSN, MS; Jill Meinert; Laura Miriel, BS; Emily P. Mitchell, MPH, MBA; Carrie Shade, BA; Jacqueline Smith, AA; Alice Sternberg, ScM; Annette Wagoner; Laura A. Wilson, ScM; Tinsay Woreta, MD, MPH; Katherine P. Yates, ScM

## Abbreviations

NAFLD: Nonalcoholic Fatty Liver Disease
PNPLA3: patatin like phospholipase domain containing 3
TM6SF2: transmembrane 6 superfamily member 2
NASH: nonalcoholic steatohepatitis
NASH CRN: NASH Clinical Research Network
GWAS: genome-wide association studies
SNPs: single nucleotide polymorphisms
NCAN: neurocan
GCKR: glucokinase regulator
LYPLAL1: lysophospholipase like 1
MBOAT7: membrane bound O-acyltransferase domain containing 7
ENPP1: ectonucleotide pyrophosphatase/phosphodiesterase 1
IRS1: insulin receptor substrate 1
SOD2: mitochondrial superoxide dismutase 2
KLF6: Kruppel-like factor 6
LPIN1: Lipin-1
UCP2: uncoupling protein 2
WES: whole-exome sequencing
HSD17B13: hydroxysteroid 17-beta dehydrogenase
MARC1: mitochondrial amidoxime reducing component 1
BMI: body mass index
H&E: hematoxylin and eosin stain
NAS: NAFLD Activity Score
PCs: Principal components
ATAV: Analysis Tool for Annotated Variants
FET: Fisher’s exact test
MAF: minor allele frequency
ExAC: Exome Aggregation Consortium
LOF: loss-of-function
SD: standard deviation
OR: odds ratio

