## Supplementary Tables 1 and 2; Supplementary Figure 1. for "Investigating the Relationship Between Rare Genetic Variants and Fibrosis in Pediatric Nonalcoholic Fatty Liver Disease"

**Supplementary Materials**

Supplementary Table 1. Gene-based variant framework collapsing models.

Supplementary Table 2A-D. Summary of Pediatric NAFLD Advanced Fibrosis Analysis Stratified by *PNPLA3* I148M Carrier Status.

Supplementary Figure 1A. Scatterplot of PC1 vs PC2 From EIGENSTRAT Analysis of Pediatric NAFLD Cases and Population Controls.

Supplementary Figure 1B. Scatterplot of PC1 vs PC2 From EIGENSTRAT Analysis of Pediatric NAFLD Cases Only.

**Supplementary Tables:**

**Supplementary Table 1. Gene-based variant framework collapsing models.**

| **Standard Variant Models** | **LOO-MAF** | **EVS MAF** | **ExAC MAF** | **Functional Categories** |
| --- | --- | --- | --- | --- |
| **Loss-of-function (LoF)** | 0.1% | 0.1% | 0.1% | LoF |
| **Rare damaging** | 0.05% | 0.01% | 0.005% | LoF, inframe indels, PolyPhen-2 "probably" |
| **Ultra-rare damaging** | 0.05% | 0% | 0% | LoF, inframe indels, PolyPhen-2 "probably" |
| **MAF<1% damaging** | 1.0% | 1.0% | 1.0% | LoF, inframe indels, PolyPhen-2 "probably" |
| **MAF<1% all functional** | 1.0% | 1.0% | 1.0% | LoF, inframe indels, all missense |
| **Recessive (1% MAF)** | 1% | 1% | 1% | LoF, inframe indels, all missense |
| **Neutral (null negative control model)** | 0.05% | 0.01% | 0.005% | Synonymous |
| **Additional models for Case vs Pop Control Analyses:** |  |  |  |  |
| **MAF<0.1% damaging** | 0.1% | 0.1% | 0.1% | LoF, inframe indels, PolyPhen-2 "probably" |
| **MAF<0.1% all functional** | 0.1% | 0.1% | 0.1% | LoF, inframe indels, all missense |

*LOO= Leave-one-out; MAF=Minor allele frequency; LoF=Loss-of-function; EVS=Exome Variant Server; ExAC=Exome Aggregation Consortium; Gnomad=Genome Aggregation Database.

**Supplementary Table 2A. Summary of Pediatric NAFLD Advanced Fibrosis Analysis Stratified by *PNPLA3* I148M Carrier Status.** Results of Collapsing Analysis of Advanced Fibrosis Phenotype Restricted to Patients with One or More PNPLA3 148M Allele (CG or GG genotype).

| **Model** | **Top Gene** | **Qualified Case Freq (AdvFib)** | **Qualified Control Freq**  **(NoFib)** | **Direction of Enrichment** | **P (FET)*** | **Lambda** |
| --- | --- | --- | --- | --- | --- | --- |
| Functional Dominant MAF<1% | *MUC5B* | 0.0102 | 0.280 | control | 1.29E-3 | 1.02 |
| Functional Recessive MAF<1% | *CCDC22* | 0.056 | 0 | case | 0.029 | 1.01 |
| Functional Dominant MAF<0.1% | *CACNA2D4* | 0 | 0.080 | control | 2.55E-3 | 1.01 |
| Damaging Dominant MAF<1% | *MYH15* | 0.075 | 0 | case | 7.03E-3 | 1.01 |
| Damaging Dominant MAF<0.1% | *PPFIBP2* | 0 | 0.060 | control | 0.012 | 0.99 |
| Rare Damaging | *PPFIBP2* | 0 | 0.040 | control | 0.053 | 0.99 |
| UltraRare Damaging | *AR* | 0.047 | 0 | case | 0.060 | 1.03 |
| LoF MAF<1% | *CXCR1* | 0 | 0.040 | control | 0.053 | 0.99 |
| LoF NoMAF threshold | *LEPREL2* | 0.486 | 0.73 | control | 3.78E-04 | 1.06 |
| Synonymous  Negative Control | *COL2A1* | 0.084 | 0 | case | 3.40E-3 | 0.98 |

*Bonferroni threshold for statistical significance was 2.68E-6.

**Supplementary Table 2B. Summary of Pediatric NAFLD Advanced Fibrosis Analysis Stratified by *PNPLA3* I148M Carrier Status.** Results of Collapsing Analysis of Advanced Fibrosis Phenotype Restricted to Patients with No *PNPLA3* 148M Alleles (CC genotype).

| **Model** | **Top Gene** | **Qualified Case Freq (AdvFib)** | **Qualified Control Freq**  **(NoFib)** | **Direction of Enrichment** | **P (FET)** | **Lambda** |
| --- | --- | --- | --- | --- | --- | --- |
| Functional Dominant MAF<1% | *BAZ2A* | 0.5 | 0 | case | 9.57E-3 | 1.01 |
| Functional Recessive MAF<1% | *GPKOW* | 0.125 | 0 | case | 0.364 | 1.06 |
| Functional Dominant MAF<0.1% | *TAS1R3* | 0.375 | 0 | case | 0.036 | 1.04 |
| Damaging Dominant MAF<1% | *COL6A2* | 0.375 | 0 | case | 0.036 | 0.99 |
| Damaging Dominant MAF<0.1% | *KCNN3* | 0.25 | 0 | case | 0.121 | 1.02 |
| Rare Damaging | *KCNN3* | 0.25 | 0 | case | 0.121 | 1.04 |
| UltraRare Damaging | *KCNN3* | 0.25 | 0 | case | 0.121 | 1.09 |
| LoF MAF<1% | *NEB* | 0.25 | 0 | case | 0.121 | 1.04 |
| LoF NoMAF threshold | *P2RX5* | 0.5 | 1 | control | 9.57E-3 | 1.07 |
| Synonymous  Negative Control | *LAMC3* | 0.375 | 0 | case | 0.036 | 1.00 |

*Bonferroni threshold for statistical significance was 2.68E-6.

**Supplementary Table 2C. Summary of Pediatric NAFLD Advanced Fibrosis Analysis Stratified by *PNPLA3* I148M Carrier Status.** Results of Common Variant (MAF>1%) Logistic Regression Analysis of Advanced Fibrosis Phenotype Restricted to Patients with One or More *PNPLA3* 148M Allele (CG or GG genotype).

| **Variant** | **Gene** | **Effect** | **OR** | **P** |
| --- | --- | --- | --- | --- |
| 13-46541673-T-A | *ZC3H13* | Glu1429Asp | 2.77 | 2.17E-4 |
| 1-248059423-C-T | *OR2W3* | Arg179Cys | 2.13 | 4.22E-4 |
| 12-58335626-A-T | *XRCC6BP1* | Ser48Cys | 2.16 | 4.91E-4 |
| 1-248039451-C-T | *TRIM58* | Thr374Met | 0.41 | 5.08E-4 |
| 19-38827960-T-C | *CATSPERG* | Leu29Pro | 0.39 | 5.52E-4 |
| 13-46629944-A-G | *CPB2* | Ile347Thr | 2.60 | 5.96E-4 |
| 16-5128817-G-A | *ALG1* | Ser267Asn | 2.82 | 6.06E-4 |
| 7-158536345-A-G | *ESYT2* | Ser584Pro | 3.62 | 6.51E-4 |
| 19-44055726-T-C | *XRCC1* | Gln399Arg | 2.40 | 7.76E-4 |
| 1-205492679-G-A | *CDK18* | Gly67Arg | 0.32 | 7.85E-4 |

*Adjusted for ancestry PCA covariates. Threshold for statistical significance after Bonferroni adjustment was p<1.77E-6. Genomic inflation factor (λ)=1.0

**Supplementary Table 2D. Summary of Pediatric NAFLD Advanced Fibrosis Analysis Stratified by *PNPLA3* I148M Carrier Status.** Results of Common Variant (MAF>1%) Logistic Regression Analysis of Advanced Fibrosis Phenotype Restricted to Patients with No *PNPLA3* 148M Alleles (CC genotype).

| **Variant** | **Gene** | **Effect** | **OR** | **P** |
| --- | --- | --- | --- | --- |
| 10-102684380-C-A | *FAM178A* | Ser541Tyr | 0.008 | 0.022 |
| 19-36168914-T-C | *UPK1A* | Met257Thr | 0.02 | 0.023 |
| 17-48432324-G-C | *XYLT2* | Arg305Thr | 0.01 | 0.024 |
| 17-81043175-G-T | *METRNL* | Ala178Ser | 273.1 | 0.025 |
| 3-42772038-A-T | *CCDC13* | Ser547Thr | 73.7 | 0.027 |
| 13-103449202-T-C | *KDELC1* | Ile114Val | 0.01 | 0.027 |
| 17-48452978-A-AAGC | *EME1* | Lys137_Pro138insGln | 111.5 | 0.028 |
| 19-36224705-A-G | *KMT2B* | Asp2364Gly | 0.02 | 0.028 |
| 2-100915772-A-G | *LONRF2* | Leu426Pro | 0.02 | 0.030 |
| 17-48456193-T-C | *EME1* | Ile350Thr | 0.01 | 0.030 |

*Adjusted for ancestry PCA covariates. Threshold for statistical significance after Bonferroni adjustment was p<1.77E-6. Genomic inflation factor (λ)=1.0.

**Supplementary Figures:**

**Supplementary Figure 1A. Scatterplot of PC1 vs PC2 From EIGENSTRAT Analysis of Pediatric NAFLD Cases and Population Controls**. Red points indicate scores for pediatric NAFLD cases, blue points indicate scores for population control subjects. Points indicate subjects score on the first and second principal components.

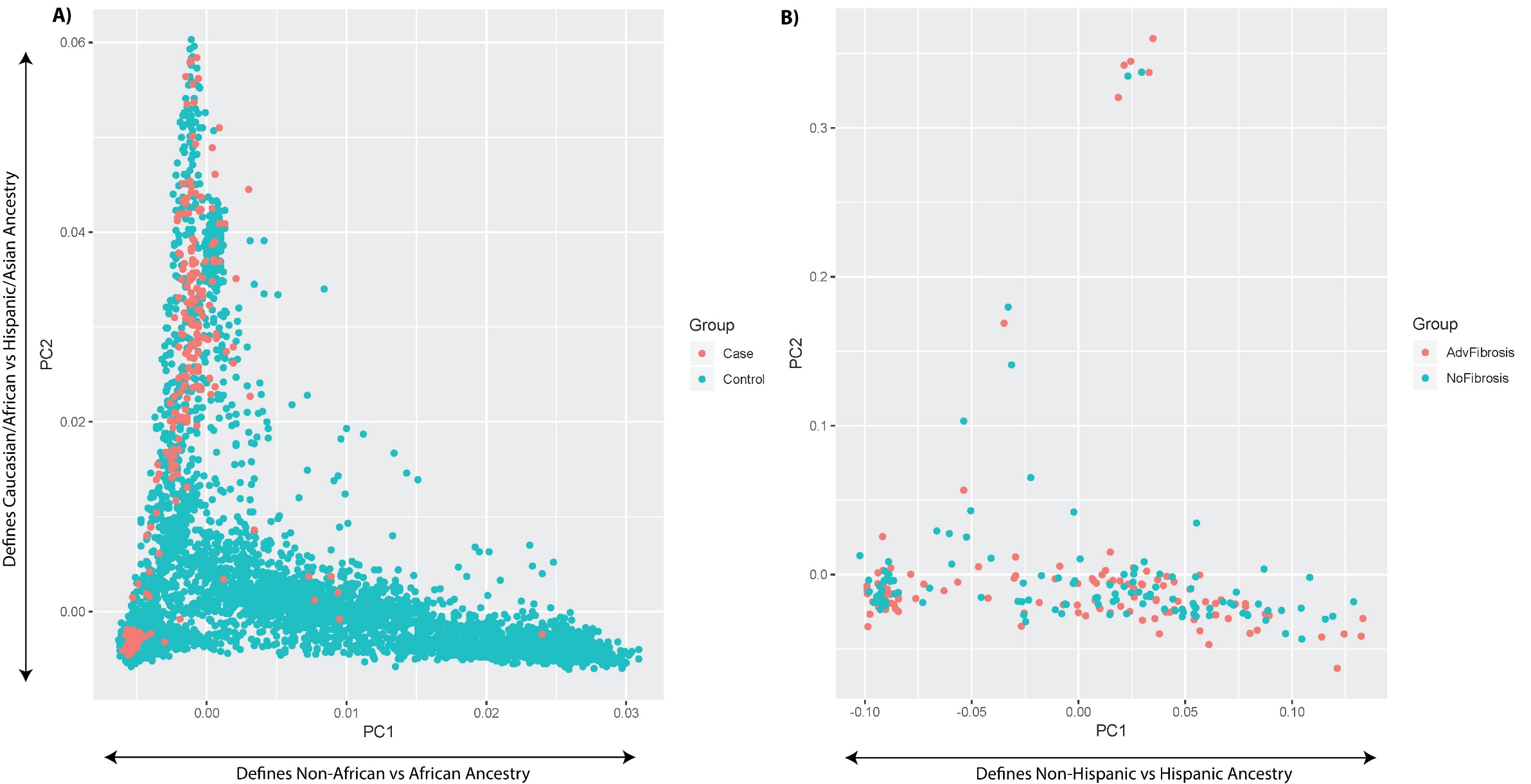

**Supplementary Figure 1B. Scatterplot of PC1 vs PC2 From EIGENSTRAT Analysis of Pediatric NAFLD Cases Only.** Red points indicate scores for pediatric NAFLD patients with advanced fibrosis (cases), while blue points indicate scores for pediatric NAFLD patients without fibrosis (controls). Points indicate the subjects score on the first and second principal components.

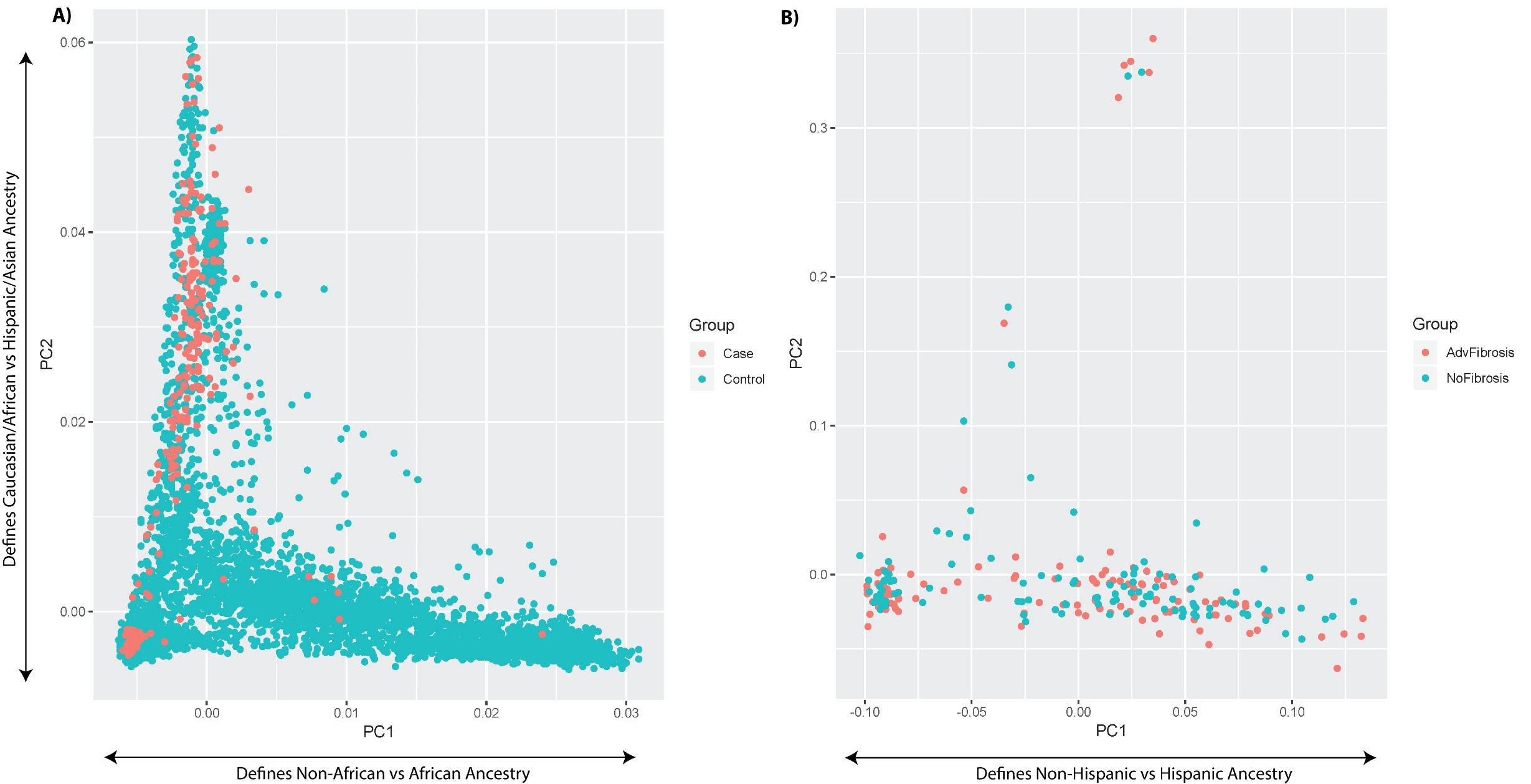
